# Machine Learning-Based Non-Invasive Diagnosis of Anemia in Children Using Palm Image Analysis

**DOI:** 10.64898/2026.01.27.26344955

**Authors:** Fatemeh Keneshlou, Mohammad Rabiee, Mo K. Delos

## Abstract

Anemia, particularly iron-deficiency anemia, is a critical global health concern, with a high prevalence among children under six years of age. Early and non-invasive detection can significantly improve health outcomes. This study proposes a computer vision and machine learning framework for anemia screening and hemoglobin (Hb) level prediction using palmar images from pediatric subjects. The region of interest (palm) was segmented using a U-Net model, achieving a Dice coefficient of 0.96. Images were processed across RGB, CIELab, and HSV color spaces to extract key color features, including red fraction, erythema index, and normalized a-component. For anemia classification, multiple machine learning models were evaluated, with Logistic Regression, Gradient Boosting, and a custom Convolutional Neural Network (CNN) achieving the highest test accuracies of approximately 94.5% and 95.53%, respectively. For hemoglobin regression, a Random Forest model in the CIELab color space achieved a coefficient of determination (R^2^) of 0.95. The Pearson correlation coefficient in the Lab color space was 0.98 for the Random Forest algorithm and 0.94 for the Linear Regression algorithm. The analysis, supported by SHAP values, identified red-related color features as the most significant predictors. The model demonstrated robust performance across different skin tones, with particularly high accuracy (R^2^ = 0.9926) in darker-skinned individuals, who constituted the majority of the studied Iranian population. The results confirm that pallor analysis of palmar images using artificial intelligence techniques offers a reliable, non-invasive, and effective tool for pediatric anemia screening and hemoglobin estimation, with strong potential for point-of-care applications.

## 1. Introduction

Anemia is a common hematologic disorder characterized by a deficiency in red blood cells or hemoglobin. Hemoglobin within red blood cells binds oxygen and transports it through capillaries to tissues throughout the body. Since all human cells depend on oxygen for survival, a deficiency leads to hypoxia and a wide range of health problems. According to the World Health Organization, 24.8% of the global population and 42% of children under six are affected by anemia, making it a significant global health issue [1].

Hemoglobin levels below 11 g/L in children, 13 g/L in men, and 12 g/L in non-pregnant women indicate anemia, while in adults, mild anemia corresponds to 9 g/L and severe anemia to 7 g/L. Iron-deficiency anemia is one of the most prevalent types, particularly in children under five. Affected children may suffer from short-and long-term health issues, including impaired growth, reduced cognition and attention, psychological disorders, and intellectual disability [2]

Anemia manifests when blood cannot meet cellular oxygen demands, and patients often require blood transfusions and repeated monitoring based on hemoglobin levels. Complete blood count (CBC), including red blood cells, white blood cells, hematocrit, hemoglobin, and platelets, is typically the first test performed to diagnose hematologic disorders. Low hemoglobin and hematocrit values generally suffice to identify iron-deficiency anemia; however, additional parameters, such as mean corpuscular volume (MCV) and mean corpuscular hemoglobin (MCH), are also considered. Red cell distribution width (RDW) is another important metric, reflecting variation in red blood cell size; high RDW values may indicate nutritional deficiencies (iron, folate, or vitamin B12) or macro/microcytic anemia[3].

Non-invasive anemia diagnosis can be achieved by analyzing patient images of specific body regions. Color, perceived through light reflection and human vision, can be quantified mathematically in color spaces, which provide coordinate-based representations of colors. Commonly used three-dimensional color spaces, aligned with human trichromatic vision, include RGB, CIELab, and HSV. The RGB color space is widely used for digital imaging, while CIELab (L = lightness, a = green-red axis, b = blue-yellow axis) is designed to match human color perception and is commonly applied in medical colorimetry. In Lab space, color differences can be quantified and compared across samples [4].

The HSV color space describes color in terms of hue, saturation, and value (brightness), facilitating image-processing algorithms by separating chromatic content from intensity. Grayscale imaging, another common color space, represents intensity without color information and is used in black-and-white images.

Early detection of anemia can improve patient outcomes, and machine learning (ML) provides a powerful tool for non-invasive diagnosis. In this study, patient images were collected under standardized conditions, ensuring uniform lighting and patient positioning, and excluding individuals with skin conditions such as jaundice or bruising. Regions of interest for anemia detection included the palmar surface, lower conjunctiva, tongue, oral mucosa near the lower lip, and fingertips. Images underwent preprocessing steps such as resizing, white-balance correction, and histogram equalization to standardize color and intensity [1].

The main objective of this study is to predict anemia in children under six years of age. As previously mentioned, this disease is associated with social, economic, hygienic, and dietary factors, and its prevalence is higher in developing countries. Therefore, there is a strong need for screening methods that enable early diagnosis in this sensitive age group. Early detection and effective treatment play a crucial role in improving the health-related quality of life of both children and the community. Accordingly, the present study proposes an effective approach for children and individuals who require frequent blood tests, which can also be made available to the general public for home-based examination and monitoring. This study employs artificial intelligence techniques, including machine learning and data mining, to process data collected from a specialized children’s hospital. It aims to investigate and identify the most appropriate methods for the diagnosis and prediction of anemia, and Hb prediction to be used as complementary diagnostic tools for physicians and patients.

## 2. Literature Review

### 2.1. Related Research on Anemia Detection via Colorimetry and Image Processing

data mining and studies on anemia have extensively utilized optical colorimetric methods, and this section is dedicated to reviewing the most recent and significant studies in this area. A common theme across these articles is the assessment of pallor in the skin and related regions, which is evaluated by measuring color indices. Skin and exposed tissue pallor can occur due to low hemoglobin levels in the bloodstream, and this phenomenon is particularly visible in areas where blood vessels are close to the surface.

The selection of regions for imaging is an important consideration in disease diagnosis. For anemia, suitable areas for imaging and subsequent image processing include: the nail bed, sclera (white of the eye), lower conjunctiva (inner membrane of the lower eyelid), oral mucosa near the lower lip, palm, tongue, and fingertips [1]. The sclera plays a role in anemia detection similar to its role in jaundice, due to the network of blood vessels and the overlaying tear layer, which provides high light reflectance and enables color measurement [5].

Data processing is performed in the CIELab color space, specifically by measuring the a-axis values, which indicate red light reflectance (green light absorption) and correspond to higher hemoglobin concentration, using machine learning techniques. According to previous studies, the redness index, derived from the a-axis values in this color model, is a key determinant: for individuals with higher hemoglobin, the mean value is 160, whereas for individuals with lower hemoglobin, the mean value is 142. Each image is first converted to the RGB color space, and the color components are extracted. These components reflect the hemoglobin concentration in the blood, which affects the color of the skin and tissues [1].

The study by Appiahene, P., et al. collected data through imaging of the palm, nails, and conjunctiva of children in Ghana as regions for anemia identification and diagnosis, with a total of 527 subjects. In this study, after the data preprocessing stage, the images were converted to the Lab color space and analyzed using Convolutional Neural Networks (CNNs), k-Nearest Neighbors (k-NN), Naive Bayes, Support Vector Machines (SVM), and Decision Trees. The results showed that Naive Bayes achieved the highest accuracy of 99.96%, while SVM had the lowest accuracy of 96.34% for anemia detection, and CNN achieved an accuracy of 99.92% in identifying the disease. To increase the dataset size for creating a more accurate model and to better evaluate the algorithm performance, the images were rotated 90°, 180°, and 270° and also flipped horizontally, ultimately increasing the total number of images fourfold [6]. Another study by Yalçın, S.S., et al. investigated beta-thalassemia in individuals aged 2 to 32 years (mean age 14.7) with a total of 105 participants in Turkey. Pallor in the palm, conjunctiva, and buccal mucosa was evaluated for beta-thalassemia diagnosis, with reported accuracies of 93.2%, 90.9%, and 80.7%, respectively, for these regions [7]. Rizal et al. also used palm imaging and pallor analysis for anemia diagnosis, employing a convolutional neural network model. By analyzing color indices in the RGB color space, they achieved a sensitivity of 100%, a specificity of 96.43%, and an accuracy of 92.86% [8].

In the study conducted by Manino and colleagues on the nail region, it was reported that there is a direct relationship between skin tone and hemoglobin levels. Since the nail contains minimal melanin compared to other areas, it is considered a suitable region for analysis. Then, the relationship between each parameter and hemoglobin level was calculated using Pearson’s correlation coefficient. The results indicated that the higher the hemoglobin level, the greater the values of the redness index and the red color fraction in the studied regions [9].

**Fig. 1.**
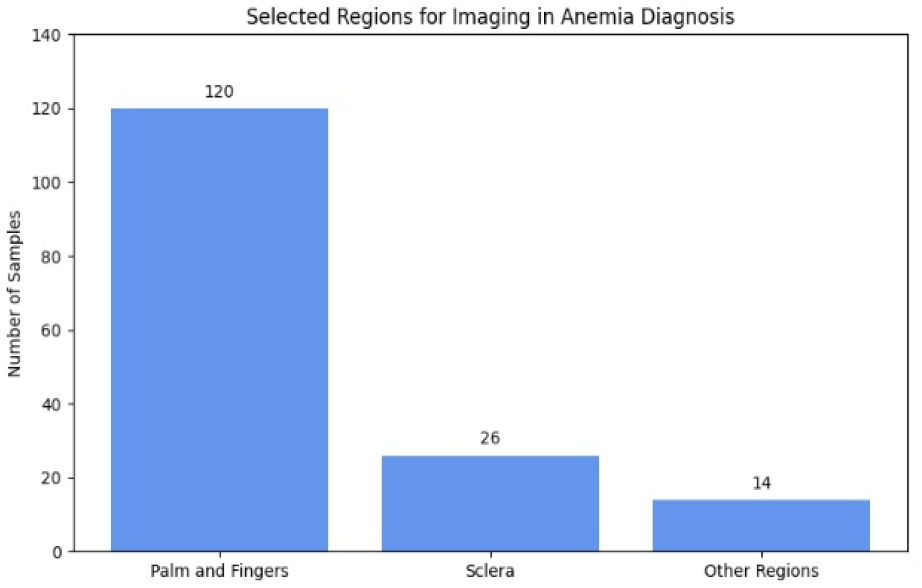
Number of published articles by region of interest (ROI) used for image-based anemia diagnosis.

Among the published studies on selecting the optimal anatomical region for anemia detection, there is ongoing debate between the conjunctival region and hand-related areas, including the palm, nail bed, and fingers. In a comprehensive study, Getaneh et al. evaluated the conjunctiva, tongue, nail, and palm regions in 574 children under five years of age in Ethiopia. Their findings indicated that the palmar region achieved higher diagnostic accuracy than the conjunctiva for anemia detection [10].

In many developed countries, patients and physicians have access to mobile health applications that enable disease screening for preliminary self-assessment and initial clinical evaluation. In the context of anemia, applications such as Anemocheck[11], HbMeter, HemaApp[12], and Sanguina allow users to monitor the condition by capturing images of specific anatomical regions. These applications provide information such as the individual’s hemoglobin level and enable the transfer of results to the relevant physician for more comprehensive evaluation. The accuracy of these applications is assessed by determining the difference and the degree of correlation between the hemoglobin values estimated by the application and those reported from the individual’s blood test.

Other diseases that can be diagnosed using colorimetry and image processing combined with machine learning include hyperbilirubinemia (jaundice)[13], skin cancer[14] and skin lesion segmentation, diabetes[15], cardiovascular and coronary artery diseases[16], COVID-19, and eye-related diseases, including iridology.

Research showed that various AI techniques, including image processing, wearable devices, and expert systems, offer remarkable accuracy and usability in diverse clinical applications. SVM is the most used algorithm, with KNN closely following. Decision Tree and ANN are also prominent. Yeruva et al. conducted a study comparing anemic and non-anemic images for anemia detection using k-NN, SVM, and decision tree classifiers.

The analysis was based on conjunctival images, while hemoglobin (Hb) values obtained from blood samples were used as auxiliary reference data. To enhance data quality, the images underwent preprocessing, including noise filtering and conversion to grayscale. Among the evaluated methods, the decision tree classifier achieved the highest accuracy (95%), outperforming SVM and k-NN, which attained accuracies of 80% and 83%, respectively [17].

**Table 1.** Summary of non-invasive anemia detection studies using image processing and machine learning techniques across different regions of interest and datasets.

| **Year & Authors** | **Participants (Age, Number, Ethnicity)** | **Region Studied** | **Image Analysis / ML Method** | **Performance Metrics** |
| --- | --- | --- | --- | --- |
| **Collings[18] et al., 2016** | 94 adults (mixed ethnicity) | Palpebral conjunctiva | Conjunctival erythema index from calibrated digital photographs | Sensitivity: 93% (training), 57% (validation); Specificity: 78% / 83% |
| **Soner[19] et al., 2018** | 710 subjects | Eye conjunctiva | CNN, SVM, Naïve Bayes, k-NN, Decision Tree; ROI extraction, color space conversion, segmentation, intensity analysis | Accuracy: CNN 99.12%; SVM 95.4% |
| **Bauskar[20] et al., 2019** | 99 subjects, India | Eye conjunctiva | Mean pixel intensity (R & G channels) + SVM | Accuracy: ~93% |
| **Jain[21] et al., 2019** | 265 patients + 72 healthy subjects | Eye conjunctiva | SVM, ANN, k-NN, FFNN, Elman NN, Ridge Regression; OpenCV-based features | MAE: 0.99 g/dL (train), 1.34 g/dL (test); RMSE: 1.29–1.72 g/dL; Pearson r ≈ 0.70 |
| **Jain et al., 2020 (MANIT Bhopal)** | 99 subjects, India | Eye conjunctiva | SVM; manual ROI selection; CIELAB color space | Accuracy: 93%; Sensitivity & Specificity reported |
| **Uma[22] et al., 2021** | 100 adults, India | Eye conjunctiva | ML classifiers (SVM, k-NN, Naïve Bayes) using RGB color features | Sensitivity: up to 96%; Specificity: up to 94% |
| **Womiz[23] et al., 2022** | 43 children, Ghana | Conjunctiva, lips, sclera | Multiple ML classifiers | Sensitivity: 90%; Specificity: 93% |
| **Womiz et al., 2022 (Ghana Dataset)** | 527 images (augmented to 2,635), children, Ghana | Eye conjunctiva | Naïve Bayes, k-NN, CNN, Decision Tree, SVM; preprocessing & augmentation | Accuracy: NB 99.96%, k-NN 99.92%, CNN 99.92%, DT 97.32%, SVM 94.94% |
| **Asare[1] et al., 2023** | Children, Ghana (multi-site dataset) | Fingernails, palm, eye conjunctiva | ML classifiers (Naïve Bayes, k-NN, SVM, Decision Tree); RGB/Lab color features | Accuracy: up to ~99%; palm & fingernails outperformed conjunctiva |
| **BioData Mining Study[6], 2023** | 527 palm images (augmented to 2,635) | Palmar region | CNN, k-NN, Naïve Bayes, SVM, Decision Tree | Accuracy: NB 99.96%; CNN 99.92% |
| **Uma[22] et al., 2025 (Eye-Defy Dataset)** | 218 images | Eye conjunctiva | Optimized SVM with SMOTE + ML classifiers | Accuracy: ~94% |
| **Ramos[24] et al., 2025** | 651 images (273 anemia, 378 non-anemia) | Conjunctiva & sclera | Vision Transformer (ViT) with attention-based explainability | Accuracy: ~98.47%; high precision & recall |

Other methods like Naïve Bayes and Random Forest are used less frequently. The study emphasizes the importance of large datasets and image augmentation to enhance machine learning performance, suggesting that SVM, KNN, and Decision Tree are particularly effective for non-invasive anemia detection. Also from the studies, CNNs demonstrated high accuracy in image-based anemia detection, Also and wearable devices provided user-friendly solutions for frequent screening. Despite these advancements, challenges such as data quality, class imbalance, and computational complexity were noted, which could impact model generalizability [25].

Apiah et al. demonstrated that the palmar region is a reliable and effective anatomical site for non-invasive anemia detection, achieving superior performance compared to earlier approaches based on conjunctival analysis. Their proposed method offers a simpler and safer alternative, particularly for pediatric populations. The authors further suggested that diagnostic accuracy could be enhanced in future studies by integrating multimodal image data from multiple body regions, such as the palm, conjunctiva, and fingernails [24].

Among the parameters investigated for anemia diagnosis are the red color fraction and the mean red color intensity, which are used in a relative manner to compare color pixels within the selected color space. In one study, red color intensity demonstrated the highest coefficient of determination (R^2^ = 81%) and the strongest correlation with hemoglobin concentration (r = 93%). Furthermore, in the study by Mekar Anggraeni et al., the diagnostic threshold was determined using a regression equation relating red color intensity to hemoglobin concentration, with a reported cutoff value of 3.79 g/dL [26].

## 3. Methodology

This chapter presents the research methodology, including data collection methods, data analysis procedures, and feature extraction techniques. The main steps involve preprocessing, processing, and evaluating the results of each model.

### 3.1. Research Statistical Population and Data Collection

The statistical population of this study was selected from the Children’s Medical Center Hospital in Tehran, under the supervision of qualified physicians and institutional authorities. The study was conducted from July to November 2025. Ethical approval was obtained from the Research Ethics Committee of the Vice-Chancellor for Research and Technology at Tehran University of Medical Sciences, and all patients provided written informed consent. Images were collected from 178 children under the age of 7, focusing on the palm area as the region of interest (ROI). Four samples were excluded due to unavailable test results, leaving 174 samples for final analysis. The study focused on classifying participants as anemic (Hb < 11 g/dL) or non-anemic (Hb ≥ 11 g/dL).

Patient data—including medical history, condition, and consent forms—was recorded. A standardized imaging protocol was developed to ensure uniform conditions for comparing healthy and patient samples.

- Camera setup: Nikon 7500D in automatic mode, 30 cm from the palm.
- Lighting: White fluorescent lamps, controlled to maintain uniform brightness, independent of outside light conditions.
- Image adjustments: White balance correction, contrast adjustment, and color standardization using calibration cards.
- Storage: Images captured in raw format (NEF), saved with patient ID, sample number, and sampling date.

After imaging, complete blood count (CBC) tests were performed. Samples were classified as non-anemic (150 samples) or anemic due to iron deficiency (24 samples) based on hemoglobin, MCV, MCH, RDW-CV, ferritin, iron, and TIBC levels. Samples with other types of anemia (e.g., beta-thalassemia, aplastic anemia, sickle cell anemia) were excluded.

**Fig. 2.**
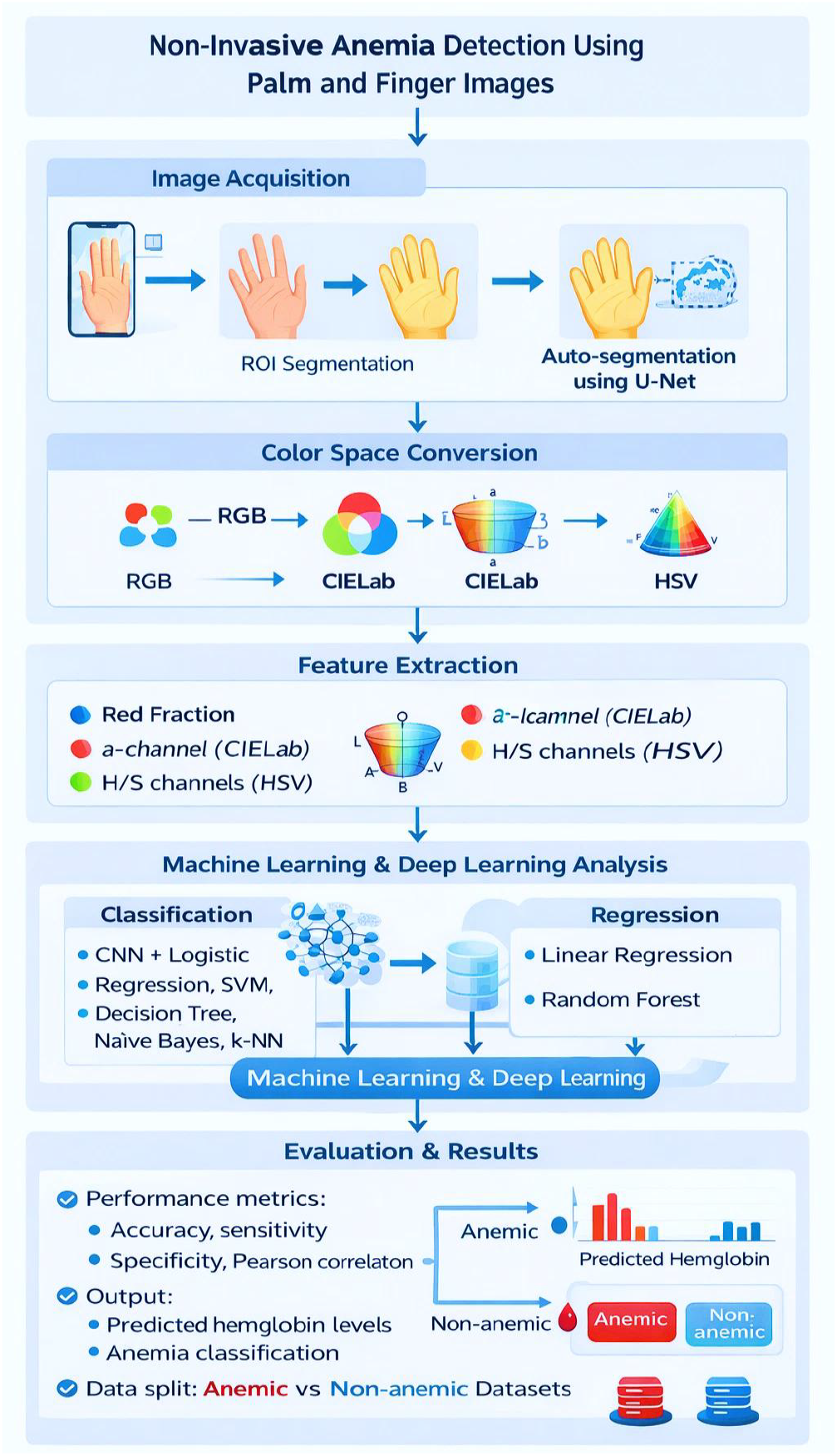
AI-Based Framework for Non-Invasive Anemia Detection from Palm and Finger Images.

### 3.2. Preprocessing

In the preprocessing stage, a U-Net model was employed for ROI segmentation instead of traditional thresholding methods. After auto-segmentation, all images were resized to uniform dimensions (224 × 224 pixels) and their white balance was adjusted. This standardization, performed using a white balance algorithm and a color calibration card, ensured that the skin tones of all samples were placed within a defined and consistent range. By default, images were converted to the RGB color space, and for subsequent processing, they were transformed into the CIELab and HSV color spaces. For each ROI, a histogram was generated to represent the three-channel color distribution in the Lab color space. To increase the variability and size of the training dataset while preserving the mean intensity values of the regions of interest (ROIs), several data augmentation techniques were applied. These included rotations of 90° and 270°, vertical and horizontal flipping, as well as random horizontal flipping, random rotations of up to ±4% of a full rotation, and random zooming along both height and width with scaling factors ranging from −5% to +5%. Finally, the images were randomly translated along both height and width axes by up to 5% of the respective dimensions. These augmentations effectively expanded the dataset and improved model generalization.

#### 3.2.1. Feature Extraction in Different Color Spaces (RGB, CIELab, HSV)

For each region of interest (ROI), the color distribution was represented as a matrix corresponding to the three RGB channels. The pixel intensity values in each channel were sorted in ascending order, and specific percentile values (5th, 25th, 50th, 75th, and 95th) were extracted to characterize the distribution. This resulted in five features per channel and a total of 15 features for the RGB color space. In addition to these percentile-based features, several primary color-based markers were extracted to characterize redness or pallor within the palm region. These included the average red intensity, erythema index, red fraction, and standardized R. These parameters were used to facilitate comparative analyses between anemic and non-anemic samples.

In the Lab color space, the L-axis represents image brightness, with values ranging from black to white. The a-axis reflects the red–green component, with positive values indicating red and negative values indicating green, while the b-axis represents the blue–yellow component. Following the same procedure as for the RGB channels, the pixel values along each axis were sorted in ascending order and specific percentiles (5th, 25th, 50th, 75th, and 95th) were extracted, resulting in a total of 15 features. Additionally, the a-axis values were standardized using min–max normalization, and the average a-component was calculated to quantify the level of redness in the images.

**Table 2.** Extracted Color Features and Their Definitions Across Different Color Spaces.

| Extracted Color Variable | Definition / Calculation | Color Space |
| --- | --- | --- |
| Erythema index | $\log(R) - \log(G)$ | RGB |
| Red fraction | $R / (R + G + B)$ | RGB |
| Minimum red | Minimum value of the red channel in the ROI | RGB |
| Normalized a | $a_{\text{norm}} = (a - \min(a)) / (\max(a) - \min(a))$ | CIELab |
| Mean a | Average value of the ‘a’ channel in the ROI | CIELab |
| Minimum hue | Minimum value of the ‘hue’ channel in the ROI | HSV |
| Mean hue | Average value of the ‘hue’ channel in the ROI | HSV |

The HSV color space was also used. In addition to the 15 percentile-based features extracted for each channel, the mean and minimum values were calculated. Relevant diagrams were generated to illustrate these parameters. In this color space, the V-axis, similar to the L-axis in the Lab color space, represents image brightness and is particularly effective for distinguishing skin tones. The H and S channels were used to compare hue and color saturation relative to white.

### 3.3. Image processing

In the present study, logistic regression, linear regression, support vector machines, decision trees, Gaussian naïve Bayes, and k-nearest neighbors algorithms were employed for both regression and classification of the data to select the best model for data categorization and to evaluate the performance of these algorithms. For the data obtained from images in the RGB and Lab color spaces. Randomly, 15%/15% of the total data were assigned to the test and validation set, while the remaining samples (70%) 120 samples were used for training. A confusion matrix was then calculated to assess the performance of the supervised learning algorithms. Model evaluation was performed using accuracy, precision, specificity, and sensitivity (recall) metrics, and the best model for classification was selected.

**Fig. 3.**
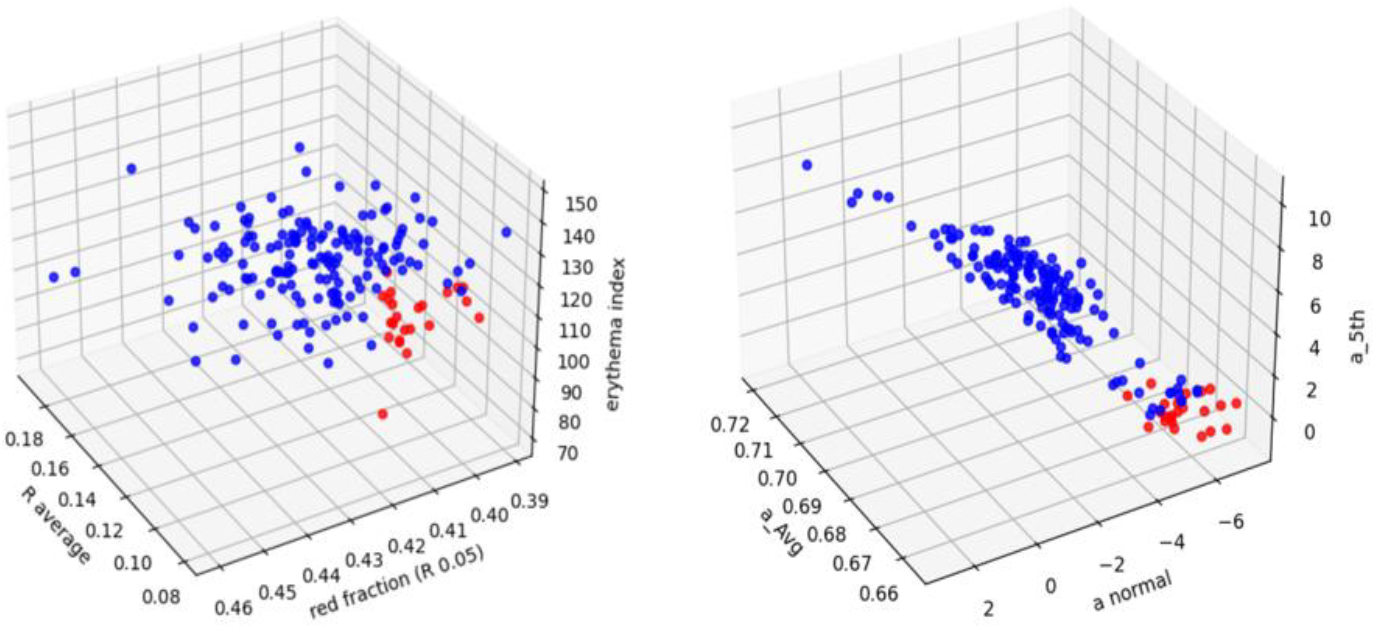
Distribution of anemic and non-anemic samples in 3D RGB and CIELab color spaces along selected color-based feature axes.

For higher accuracy to be derived, data augmentation was used to generate samples, including a 20-degree rotation, horizontal translation equal to 0.2 of the total image width, vertical translation equal to 0.2 of the total image height, a zoom range of 0.2, and shear translation of 0.2. The small size of the dataset is capable of resulting in overfitting. Due to class imbalance, the anemic class was expanded using data augmentation techniques (Photometric data augmentation), while class weights were applied during model training. This is due to the reason that during the training and testing stage, data samples that are small in number are not able to be generalized and used for hyperparameter models. The data were also used to investigate the relationship between the parameters and variables extracted from the images, the degree of pallor in the region of interest, and Hb levels. This allowed identification of the color parameters that most significantly influenced Hb prediction and data classification. In predicting Hb, the independent variable was the actual Hb value, while the dependent variables in the RGB color space included the 5th percentile red fraction, redness index, and mean red value. In the Lab color space, the dependent variables included the normalized a component and its mean. In the HSV space, the independent variable was the actual Hb value, and the dependent variables were the mean H and S channels. The correlation between actual and predicted Hb values was reported using the Pearson correlation coefficient.

### 3.4. Machine learning and deep learning models for anemia classification and Hb Prediction

In this study, regression and classification algorithms—including Logistic Regression, Linear Regression, Support Vector Machine (SVM), Decision Tree, Gaussian Naïve Bayes, and k-Nearest Neighbors (k-NN), and CNN—were used to identify the best model for data classification and to evaluate algorithm performance.

A custom convolutional neural network (CNN) consisting of four convolutional blocks followed by global average pooling and fully connected layers was developed for binary anemia classification. This model, which incorporates batch normalization, dropout, and global average pooling, was employed to improve training stability and reduce overfitting.

#### Summary of CNN model architecture

- Input: 224 × 224 × 3 RGB images
- Convolutional layers:
  - 32 → 64 → 128 → 256 filters
- Batch Normalization: after each convolution
- Pooling: MaxPooling + Global Average Pooling
- Regularization: Dropout (0.4)
- Output: Sigmoid activation (binary classification)

For the data obtained from images, a confusion matrix was computed to assess the performance of the supervised learning algorithms. Evaluation metrics including accuracy, precision, recall, F1-score, and sensitivity were used to evaluate the designed models, and the best model for classification was selected. A multiple linear regression model was employed for Hb prediction, and the coefficient of determination (*R*^2^) was calculated for each color space. The influence of each color parameter on the predicted hemoglobin value and the probability of each variable was also reported. To further examine prediction accuracy across different skin tones, samples were divided based on the mean L channel in the Lab space, which represents brightness, into light and dark skin tone categories. According to this criterion, higher L values in the region of interest corresponded to lighter skin tones. Based on the Fitzpatrick scale, which classifies human skin color into six categories, the skin tones of the Iranian samples in this study ranged from type II to IV, corresponding to the lightest to the darkest tones. Using this classification, the ability of the designed program to estimate Hb was evaluated across different skin tones.

## 4. Results

The results of auto-segmentation using U-Net demonstrate strong performance, achieving a best Intersection over Union (IoU) of approximately 0.935 and a best Dice coefficient of approximately 0.96. Additionally, the training loss decreased to a minimum value of approximately 0.15, indicating accurate segmentation and stable convergence of the model.

**Fig. 4.**
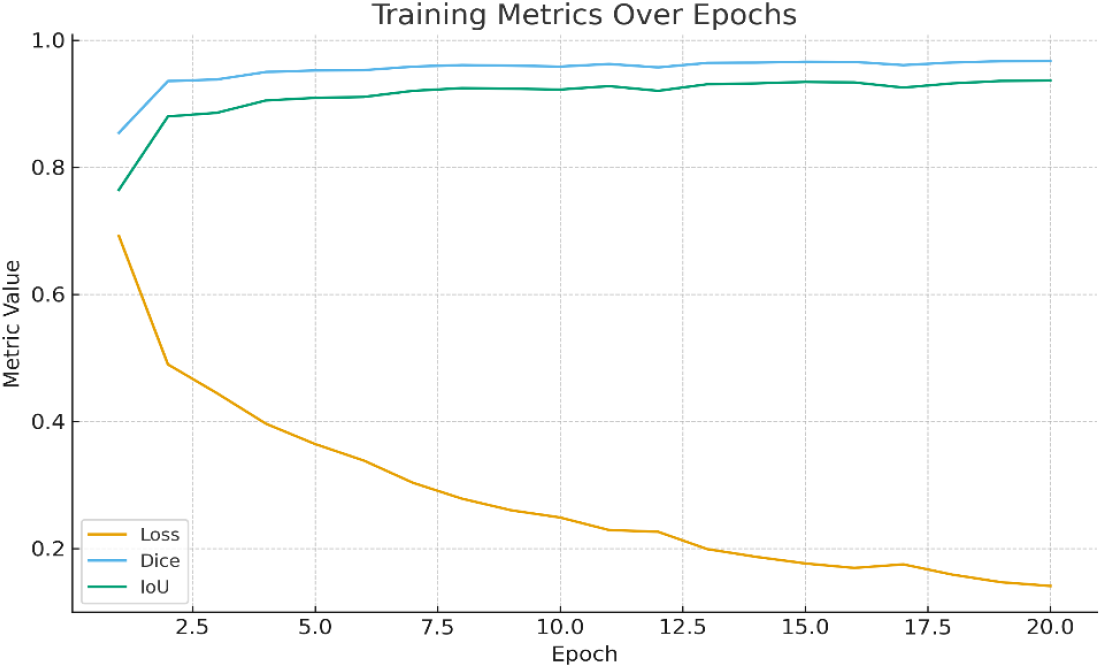
Training curves of the U-Net–based model for automatic ROI segmentation of palm and finger regions.

### 4.1. Anemia Classification

The extracted color features were used twice: first, for labeling the data based on whether the samples were healthy or anemic (classification), and second, for predicting the estimated hemoglobin level using regression models. It was shown that, in the RGB color space, the most important color features included the mean red pixel intensity of the samples, the red fraction (5%), and the redness index. In the RGB color space, as shown in Fig. 5 the performance of multiple machine learning classifiers was evaluated using ROC curves and corresponding AUC values. On the test dataset, Logistic Regression, Gradient Boosting, and AdaBoost achieved the highest classification accuracies (94.5%) with AUCs of 0.96, while k-Nearest Neighbors and Random Forest reached 93.5% (AUC 0.95). Gaussian Naïve Bayes achieved 90% accuracy with an AUC of 0.94, and Decision Tree showed the lowest performance (85%, AUC 0.86).

**Fig. 5.**
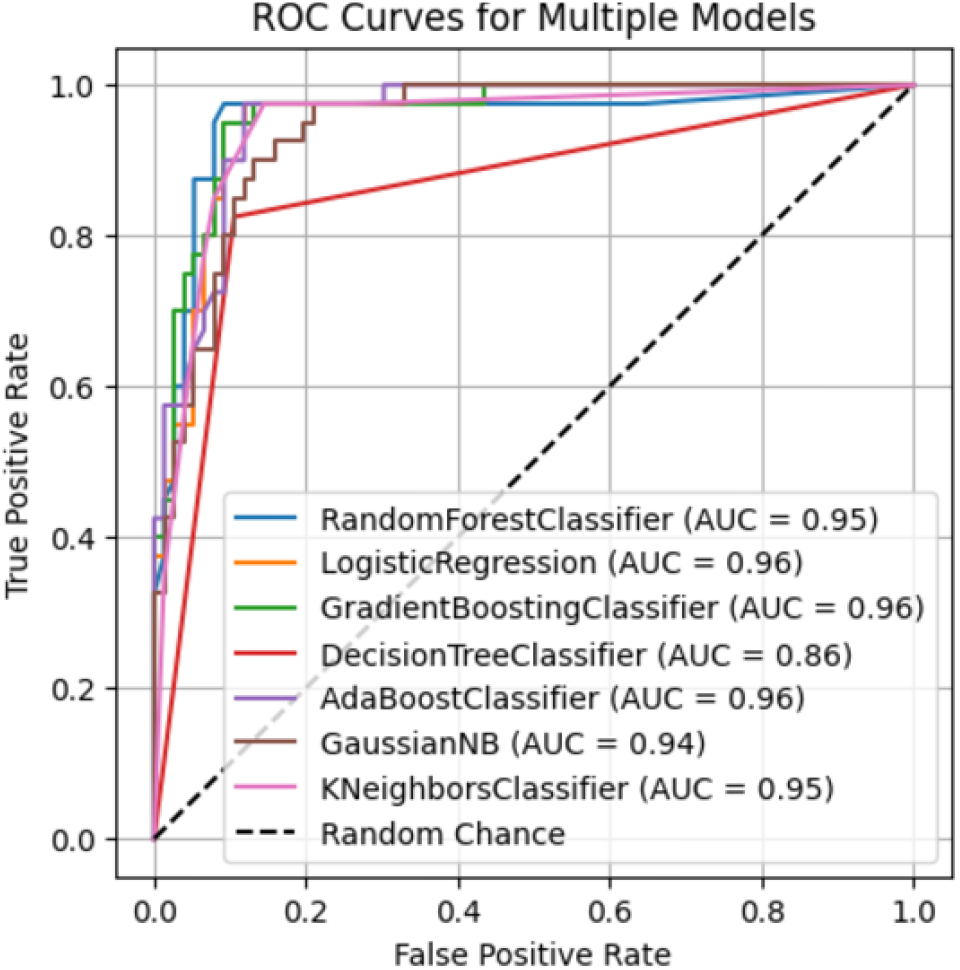
ROC curves of multiple machine learning classifiers in the RGB color space.

The model in the CIELab color space showed that the most important color features in this space were the mean of the a-axis, the normalized a-axis value, and the 5% a-axis value. Classification plots were generated based on these principal components. According to the plots and results, samples with a mean a-axis value below 115 were classified as anemic. Among the color features in the Lab color space, the most predictive feature for hemoglobin level was identified as the mean value of the a-axis, which showed the lowest probability of the null hypothesis. In this color space, a multiple linear regression model was used to fit the data. The samples were divided into two groups—light and dark skin tones—based on the L-axis value, which represents image brightness and skin tone, so that the performance of the algorithms in predicting hemoglobin could be evaluated specifically with respect to skin color.

**Table 3.** Precision, Recall, and F1-score of different classifiers for anemia classification in the RGB color space.

| Classifier | AUC | Precision | Recall | F1-score |
| --- | --- | --- | --- | --- |
| LogisticRegression | 0.96 | 0.92 | 0.93 | 0.925 |
| GradientBoosting | 0.96 | 0.92 | 0.93 | 0.925 |
| AdaBoost | 0.96 | 0.91 | 0.93 | 0.92 |
| RandomForest | 0.95 | 0.90 | 0.92 | 0.91 |
| KNeighbors | 0.95 | 0.89 | 0.91 | 0.90 |
| GaussianNB | 0.94 | 0.88 | 0.90 | 0.89 |
| DecisionTree | 0.86 | 0.78 | 0.80 | 0.79 |

Figure 5(A) shows the training and validation accuracy of the CNN model over 100 epochs. Training accuracy increases from approximately 0.90 at epoch 1 to 0.97 at epoch 100. Validation accuracy starts at approximately 0.87 and reaches 0.96–0.97 by the final epoch. Across all epochs, the difference between training and validation accuracy remains within ±0.02. In addition, Figure 5(B) presents the training and validation loss curves over the same training period. Training loss decreases from approximately 0.19 at the first epoch to 0.09 at epoch 100. Validation loss decreases from approximately 0.29 initially to 0.06 at the final epoch. During early epochs (1–30), validation loss fluctuates between 0.15 and 0.30, while after epoch 60 it remains below 0.15. The trained CNN model achieved an accuracy of 95.53% with a loss value of 0.175 on the test dataset

**Fig. 6.**
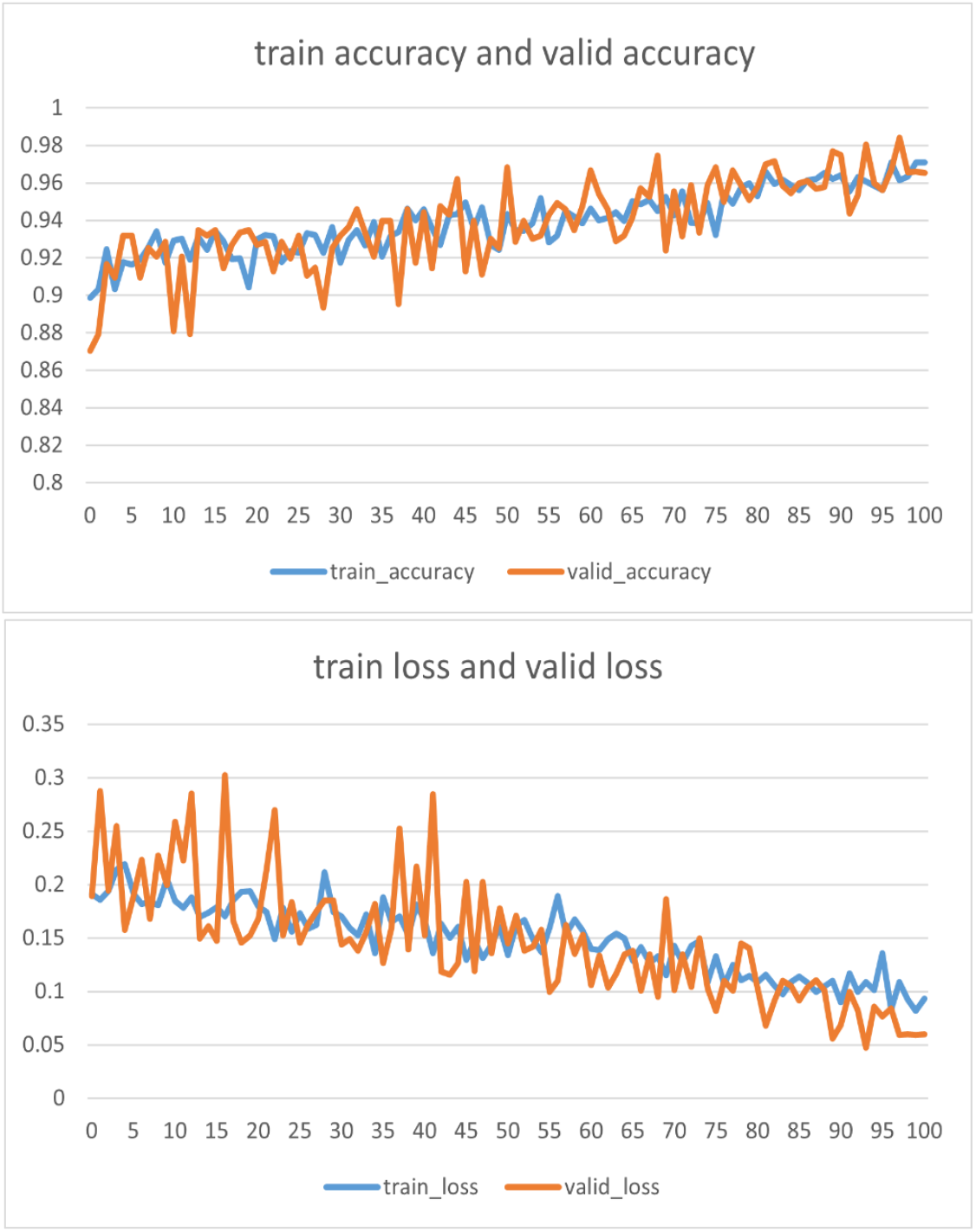
CNN training and validation performance: (A) accuracy and (B) loss over 100 epochs.

### 4.2. Hemoglobin Prediction

The results indicated that the coefficient of determination (R^2^) of the linear regression model was 85.01% for light-skinned samples, 99.26% for dark-skinned samples, and 94.37% when the model was applied without considering skin tone. Since this study was conducted on an Iranian population, the higher accuracy observed in dark-skinned samples is reasonable, given that most participants belong to the dark skin group according to the global standard classification. As shown in Fig.7, the Random Forest model achieved an R^2^ of 0.95 in the Lab color space. Additionally, the Pearson correlation coefficient for the two variables—actual hemoglobin and hemoglobin predicted by the algorithm—in the Lab color space was 0.98 for the Random Forest algorithm and 0.94 for the Linear Regression algorithm.

**Fig. 7.**
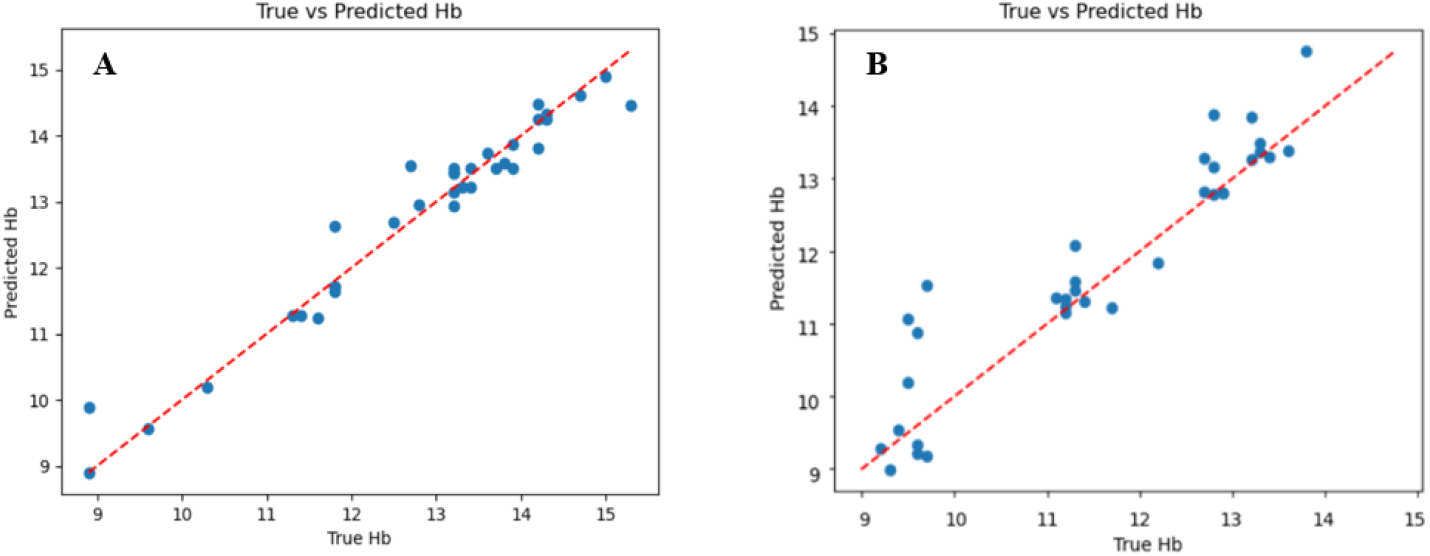
Random Forest–Based Hemoglobin Prediction: (A) Lab Color Space and (B) RGB Color Space—Actual vs. Predicted Values.

**Fig. 8.**
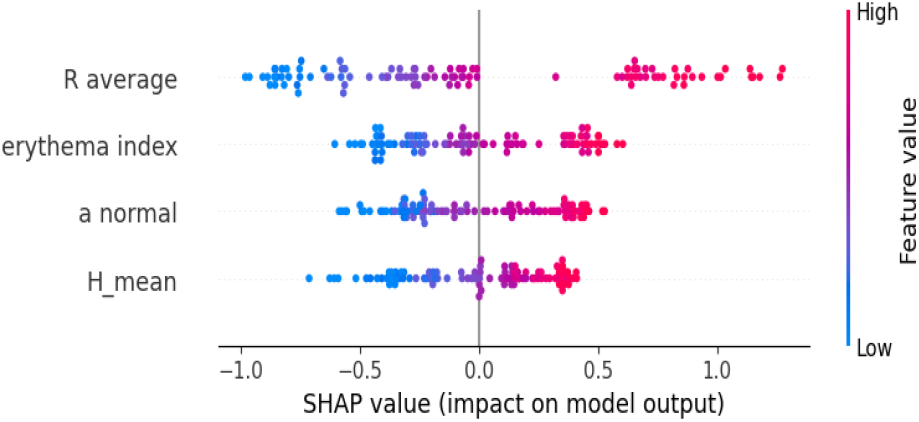
SHAP summary plot. Feature importance analysis for hemoglobin prediction using color-based parameters.

To interpret the predictions of the Random Forest Regressor, SHAP (SHapley Additive exPlanations) values were calculated. Unlike traditional feature importance, SHAP provides insight into both the magnitude and the direction of a feature’s influence on the predicted hemoglobin levels. The SHAP summary plot demonstrates that R average and erythema index are the most influential features in hemoglobin prediction. All top features exhibit a positive correlation with the model output, which means higher values of these color features consistently contribute to increased hemoglobin estimates. The concentration of blue dots on the negative side of the SHAP x-axis across all features suggests that the model is particularly sensitive to the “loss of redness,” which it uses as a primary indicator for identifying low hemoglobin (anemia) levels.

## 5. conclusion

This study investigates the comparative effectiveness of various smartphone image analysis techniques for anemia screening, focusing on a clinically vulnerable and underrepresented pediatric demographic. The primary objective was to identify the optimal analytical methods for this specific population group.

The findings of this study regarding the early and non-invasive diagnosis of anemia indicate that the best color space for comparing the extracted color features from images is the Lab color space. The most effective color metrics for classifying samples were found to be the mean a-axis value in the Lab space and the mean redness value in the RGB space. Furthermore, for hemoglobin estimation, the Random Forest and Multiple Linear Regression algorithms demonstrated the best performance. When estimating hemoglobin based on skin tone, the algorithms performed more effectively on darker-toned skin due to the larger number of data points in this group. The highest Pearson correlation coefficient between actual and model-predicted hemoglobin values was achieved in the Lab color space.

In the RGB color space, the performance of several machine learning classifiers was evaluated. Based on the ROC curve and corresponding AUC values, the classification accuracies on the test dataset These results demonstrate the strong discriminative ability of the classifiers, consistent with the high AUC values, and confirm their reliability in distinguishing anemic from non-anemic cases.

The analysis utilized an automated approach for segmenting the palmar region of interest via a U-Net model, replacing traditional manual or threshold-based segmentation. Overall, the parallel behavior of training and validation curves in both accuracy and loss plots indicates that the proposed CNN architecture achieves robust and stable learning, with strong generalization capability for anemia classification. The use of global average pooling reduces the number of trainable parameters, while dropout regularization prevents co-adaptation of neurons, contributing to the observed performance consistency across training and validation datasets.

According to SHAP plot, findings confirm that red-related color features play a dominant role in non-invasive hemoglobin estimation, highlighting their potential for clinical screening applications. Furthermore, the highest Pearson correlation coefficient between actual and model-predicted hemoglobin values was achieved using the CIELab color space.

## Supporting information

Unet-segmentation

Anemia-CNN-Classification

## Data Availability

All data produced in the present study are available upon reasonable request to the authors

## References

1. Asare, J.W., et al., Iron deficiency anemia detection using machine learning models: A comparative study of fingernails, palm and conjunctiva of the eye images. Engineering Reports, 2023. 5(11): p. e12667.

2. Akbarpour, E., et al., Anemia prevalence, severity, types, and correlates among adult women and men in a multiethnic Iranian population: the Khuzestan Comprehensive Health Study (KCHS). BMC Public Health, 2022. 22(1): p. 168.

3. Viswanath, D., et al., Red cell distribution width in the diagnosis of iron deficiency anemia. The Indian Journal of Pediatrics, 2001. 68(12): p. 1117–1119.

4. Mahmud, S., et al., Anemia detection through non-invasive analysis of lip mucosa images. Frontiers in big Data, 2023. 6: p. 1241899.

5. Dimauro, G., et al., Anaemia detection based on sclera and blood vessel colour estimation. Biomedical Signal Processing and Control, 2023. 81: p. 104489.

6. Appiahene, P., et al., Detection of iron deficiency anemia by medical images: a comparative study of machine learning algorithms. BioData mining, 2023. 16(1): p. 2.

7. Yalçin, S.S., et al., The validity of pallor as a clinical sign of anemia in cases with beta-thalassemia. The Turkish journal of pediatrics, 2007. 49(4): p. 408–412.

8. Rizal, A.S., et al., Detecting Anemia Based on Palm Images using Convolutional Neural Network. International Journal of Advanced Engineering Research and Science (IJAERS), 2022.

9. Das, S., et al. Non-invasive haemoglobin estimation by observing nail color: A PCA based approach. in 2022 IEEE 6th conference on information and communication technology (CICT). 2022. IEEE.

10. Getaneh, T., et al., The utility of pallor detecting anemia in under five years old children. Ethiopian medical journal, 2000. 38(2): p. 77–84.

11. Perez-Plazola, M.S., et al., AnemoCheck-LRS: an optimized, color-based point-of-care test to identify severe anemia in limited-resource settings. BMC medicine, 2020. 18(1): p. 337.

12. Wang, E.J., et al. HemaApp: noninvasive blood screening of hemoglobin using smartphone cameras. in Proceedings of the 2016 ACM international joint conference on pervasive and ubiquitous computing. 2016.

13. Seok, J., et al., Non-Invasive Jaundice Screening Using AI: Machine Learning Analysis of Sclera and Urine Images. Journal of Clinical Medicine, 2025. 14(9): p. 3125.

14. Yang, X., et al., A novel multi-task deep learning model for skin lesion segmentation and classification. arXiv preprint arXiv:1703.01025, 2017.

15. Zhang, B. and D. Zhang, Noninvasive diabetes mellitus detection using facial block color with a sparse representation classifier. IEEE transactions on biomedical engineering, 2013. 61(4): p. 1027–1033.

16. Kim, B.-h., et al. A proposal of heart diseases diagnosis method using analysis of face color. in 2008 International Conference on Advanced Language Processing and Web Information Technology. 2008. IEEE.

17. Yeruva, S., et al., Identification of sickle cell anemia using deep neural networks. Emerging Science Journal, 2021. 5(2): p. 200–210.

18. Collings, S., et al., Non-invasive detection of anaemia using digital photographs of the conjunctiva. PloS one, 2016. 11(4): p. e0153286.

19. Suner, S., et al., Prediction of anemia and estimation of hemoglobin concentration using a smartphone camera. PloS one, 2021. 16(7): p. e0253495.

20. Bauskar, S., P. Jain, and M. Gyanchandani, A noninvasive computerized technique to detect anemia using images of eye conjunctiva. Pattern Recognition and Image Analysis, 2019. 29(3): p. 438–446.

21. Kasiviswanathan, S., T.B. Vijayan, and S. John, Ridge regression algorithm based non-invasive anaemia screening using conjunctiva images. Journal of Ambient Intelligence and Humanized Computing, 2020: p. 1–11.

22. Uma, K., et al. Analysis of Anemia Detection from Images of Eye Conjunctiva Using Machine Learning Algorithms. in International Conference on Sustainability Innovation in Computing and Engineering (ICSICE 2024). 2025. Atlantis Press.

23. Wemyss, T.A., et al., Feasibility of smartphone colorimetry of the face as an anaemia screening tool for infants and young children in Ghana. Plos one, 2023. 18(3): p. e0281736.

24. Ramos-Soto, O., et al., Non-invasive anemia detection from conjunctiva and sclera images using vision transformer with attention map explainability. Scientific Reports, 2025. 15(1): p. 44142.

25. Mahmud, A.A., et al., AI-Driven anemia diagnosis: A review of advanced models and techniques. arXiv preprint arXiv:2510.11380, 2025.

26. Anggraeni, M. and A. Fatoni. Non-invasive self-care anemia detection during pregnancy using a smartphone camera. in IOP Conference series: materials science and engineering. 2017. IOP Publishing.

