## Supplementary material for "Machine Learning-Based Non-Invasive Diagnosis of Anemia in Children Using Palm Image Analysis": Unet-segmentation

```
import os
import json
import cv2
import torch
from torch.utils.data import Dataset, DataLoader
import albumentations as A
from albumentations.pytorch import ToTensorV2

root = "/content/drive/MyDrive/dataset"
output_json = os.path.join(root, "dataset.json")
data = {"dataset": []}

for fname in os.listdir(root):
    if not fname.lower().endswith('.jpg'):
        continue

    index = os.path.splitext(fname)[0]
    mask_name = f"Mask{index}.png"
    img_path  = os.path.join(root, fname)
    mask_path = os.path.join(root, mask_name)

    if os.path.exists(mask_path):
        data["dataset"].append({
            "image": img_path,
            "mask": mask_path
        })
        print(f"Matched: {fname} ↔ {mask_name}")
    else:
        print(f"no {mask_name}")

with open(output_json, "w") as f:
    json.dump(data, f, indent=4)

print("\nJSON saved:", output_json)

import cv2
import os
import albumentations as A

input_images = "/content/drive/MyDrive/dataset/images"
input_masks  = "/content/drive/MyDrive/dataset/masks"

output_images = "/content/drive/MyDrive/dataset/augmented/images"
output_masks  = "/content/drive/MyDrive/dataset/augmented/masks"

os.makedirs(output_images, exist_ok=True)
os.makedirs(output_masks, exist_ok=True)

augment = A.Compose([
    A.HorizontalFlip(p=0.5),
    A.ShiftScaleRotate(shift_limit=0.05, scale_limit=0.12,
rotate_limit=12, p=0.7),
    A.RandomBrightnessContrast(brightness_limit=0.2, contrast_limit=0.2,
p=0.4),
    A.HueSaturationValue(hue_shift_limit=5, sat_shift_limit=12,
val_shift_limit=12, p=0.3),
    A.GaussianBlur(p=0.2),
])
```

```
N = 20

total = 0

for fname in os.listdir(input_images):
    if not fname.lower().endswith((".jpg", ".png", ".jpeg")):
        continue

    index = os.path.splitext(fname)[0]

    img_path = os.path.join(input_images, fname)
    mask_path = os.path.join(input_masks, f"Mask{index}.png")

    if not os.path.exists(mask_path):
        print(f"no {mask_path}")
        continue

    img = cv2.imread(img_path)
    img = cv2.cvtColor(img, cv2.COLOR_BGR2RGB)
    mask = cv2.imread(mask_path, cv2.IMREAD_GRAYSCALE)

    for i in range(N):
        aug = augment(image=img, mask=mask)
        img_aug = aug["image"]
        mask_aug = aug["mask"]

        out_img = f"{output_images}/{index}_aug{i}.png"
        out_mask = f"{output_masks}/{index}_aug{i}.png"

        cv2.imwrite(out_img, cv2.cvtColor(img_aug, cv2.COLOR_RGB2BGR))
        cv2.imwrite(out_mask, mask_aug)

        total += 1

print(f"\n finish")
print(f" total images: {total}")
print(f" stored: {output_images}")
print("Total pairs:", len(data["dataset"]))

class PalmSegmentationDataset(Dataset):
    def __init__(self, json_path, transform=None):
        with open(json_path, "r") as f:
            self.data = json.load(f)

        self.transform = transform

    def __len__(self):
        return len(self.data)

    def __getitem__(self, idx):
        img_path = self.data[idx]["image"]
        mask_path = self.data[idx]["mask"]

        image = cv2.imread(img_path)
        image = cv2.cvtColor(image, cv2.COLOR_BGR2RGB)

        mask = cv2.imread(mask_path, cv2.IMREAD_GRAYSCALE)
```

```
        if self.transform:
            aug = self.transform(image=image, mask=mask)
            image = aug["image"]
            mask = aug["mask"]

        mask = (mask > 128).float()

        return image, mask

train_transform = A.Compose([
    A.Resize(256, 256),
    A.HorizontalFlip(p=0.5),
    A.ShiftScaleRotate(shift_limit=0.05, scale_limit=0.15,
rotate_limit=15, p=0.7),
    A.RandomBrightnessContrast(p=0.4),
    A.GaussianBlur(p=0.1),
    A.Normalize(mean=(0.5, 0.5, 0.5),
                std=(0.5, 0.5, 0.5)),
    ToTensorV2()
])

json_path = "/content/drive/MyDrive/dataset/augmented/dataset.json"

train_dataset = PalmSegmentationDataset(json_path, train_transform)
train_loader = DataLoader(train_dataset, batch_size=4, shuffle=True,
num_workers=2)

import torch.nn as nn
import torch

class DoubleConv(nn.Module):
    def __init__(self, in_channels, out_channels):
        super().__init__()
        self.net = nn.Sequential(
            nn.Conv2d(in_channels, out_channels, 3, padding=1),
            nn.BatchNorm2d(out_channels),
            nn.ReLU(inplace=True),

            nn.Conv2d(out_channels, out_channels, 3, padding=1),
            nn.BatchNorm2d(out_channels),
            nn.ReLU(inplace=True)
        )

    def forward(self, x):
        return self.net(x)

class UNet(nn.Module):
    def __init__(self):
        super().__init__()

        self.enc1 = DoubleConv(3, 64)
        self.enc2 = DoubleConv(64, 128)
        self.enc3 = DoubleConv(128, 256)
        self.enc4 = DoubleConv(256, 512)

        self.pool = nn.MaxPool2d(2)
```

```
        self.bottleneck = DoubleConv(512, 1024)

        self.up4 = nn.ConvTranspose2d(1024, 512, 2, stride=2)
        self.dec4 = DoubleConv(1024, 512)

        self.up3 = nn.ConvTranspose2d(512, 256, 2, stride=2)
        self.dec3 = DoubleConv(512, 256)

        self.up2 = nn.ConvTranspose2d(256, 128, 2, stride=2)
        self.dec2 = DoubleConv(256, 128)

        self.up1 = nn.ConvTranspose2d(128, 64, 2, stride=2)
        self.dec1 = DoubleConv(128, 64)

        self.out = nn.Conv2d(64, 1, 1)

    def forward(self, x):
        c1 = self.enc1(x)
        c2 = self.enc2(self.pool(c1))
        c3 = self.enc3(self.pool(c2))
        c4 = self.enc4(self.pool(c3))

        bottleneck = self.bottleneck(self.pool(c4))

        u4 = self.up4(bottleneck)
        u4 = torch.cat([u4, c4], dim=1)
        d4 = self.dec4(u4)

        u3 = self.up3(d4)
        u3 = torch.cat([u3, c3], dim=1)
        d3 = self.dec3(u3)

        u2 = self.up2(d3)
        u2 = torch.cat([u2, c2], dim=1)
        d2 = self.dec2(u2)

        u1 = self.up1(d2)
        u1 = torch.cat([u1, c1], dim=1)
        d1 = self.dec1(u1)

        return torch.sigmoid(self.out(d1))

import torch.nn.functional as F

def dice_loss(pred, target, smooth=1):
    pred = pred.view(-1)
    target = target.view(-1)
    intersection = (pred * target).sum()
    return 1 - (2. * intersection + smooth) / (pred.sum() + target.sum()
+ smooth)


def loss_fn(pred, target):
    bce = F.binary_cross_entropy(pred, target)
    dice = dice_loss(pred, target)
    return bce + dice

def dice_metric(pred, target, threshold=0.5, smooth=1):
```

```
    pred = (pred > threshold).float()
    target = target.float()

    intersection = (pred * target).sum(dim=(1,2,3))
    union = pred.sum(dim=(1,2,3)) + target.sum(dim=(1,2,3))

    dice = (2 * intersection + smooth) / (union + smooth)
    return dice.mean().item()


def iou_metric(pred, target, threshold=0.5, smooth=1):
    pred = (pred > threshold).float()
    target = target.float()

    intersection = (pred * target).sum(dim=(1,2,3))
    union = pred.sum(dim=(1,2,3)) + target.sum(dim=(1,2,3)) -
intersection

    iou = (intersection + smooth) / (union + smooth)
    return iou.mean().item()

device = "cuda" if torch.cuda.is_available() else "cpu"
model = UNet().to(device)

from torchsummary import summary
summary(model, (3, 256, 256))

optimizer = torch.optim.Adam(model.parameters(), lr=1e-4)

EPOCHS = 20

for epoch in range(EPOCHS):
    model.train()

    running_loss = 0
    running_dice = 0
    running_iou  = 0

    for imgs, masks in train_loader:
        imgs = imgs.to(device)
        masks = masks.to(device).unsqueeze(1)   # ensure shape: (B,1,H,W)

        preds = model(imgs)

        # -------- Loss --------
        loss = loss_fn(preds, masks)

        optimizer.zero_grad()
        loss.backward()
        optimizer.step()

        running_loss += loss.item()

        # -------- Metrics --------
        with torch.no_grad():
            running_dice += dice_metric(preds, masks)
            running_iou  += iou_metric(preds, masks)
```

```
    epoch_loss = running_loss / len(train_loader)
    epoch_dice = running_dice / len(train_loader)
    epoch_iou  = running_iou  / len(train_loader)

    print(f"Epoch {epoch+1}/{EPOCHS}  |  "
          f"Loss: {epoch_loss:.4f}  |  "
          f"Dice: {epoch_dice:.4f}  |  "
          f"IoU: {epoch_iou:.4f}")

torch.save(model.state_dict(), "best_unet_model.pth")
model = UNet().to(device)
model.load_state_dict(torch.load("best_unet_model.pth",
map_location=device))
model.eval()

import torch
import cv2
import numpy as np
import matplotlib.pyplot as plt

def predict_single_image(model, image_path):
    model.eval()

    img = cv2.imread(image_path)
    img = cv2.cvtColor(img, cv2.COLOR_BGR2RGB)

    img_resized = cv2.resize(img, (256, 256))

    tensor = torch.from_numpy(img_resized.transpose(2,0,1)).float() /
255.0
    tensor = tensor.unsqueeze(0).to(device)

    with torch.no_grad():
        pred = model(tensor)
        pred = torch.sigmoid(pred)
        mask = (pred.cpu().numpy()[0,0] > 0.5).astype(np.uint8)

    return img, img_resized, mask

import torch
import matplotlib.pyplot as plt

device = "cuda" if torch.cuda.is_available() else "cpu"
model = UNet().to(device)

imgs, masks = next(iter(train_loader))
imgs = imgs.to(device)
masks = masks.to(device).unsqueeze(1)

model.eval()
with torch.no_grad():
    preds = model(imgs)
    preds = torch.sigmoid(preds)


img = imgs[0].cpu().permute(1, 2, 0).numpy()
gt = masks[0].cpu().squeeze().numpy()
pr = preds[0].cpu().squeeze().numpy()
```

```
plt.figure(figsize=(12, 4))

plt.subplot(1, 3, 1)
plt.imshow(img)
plt.title("Input Image")
plt.axis("off")

plt.subplot(1, 3, 2)
plt.imshow(gt, cmap= 'gray')
plt.title("Ground Truth Mask")
plt.axis("off")

plt.subplot(1, 3, 3)
plt.imshow(pr)
plt.title("Predicted Mask")
plt.axis("off")

plt.tight_layout()
plt.show()
```

@Author : Fatemeh_Kn
