## Supplementary material for "Machine Learning-Based Non-Invasive Diagnosis of Anemia in Children Using Palm Image Analysis": Anemia-CNN-Classification

```
from sklearn.ensemble import RandomForestClassifier
import cv2 as cv
import numpy as np
import os
import matplotlib.pyplot as plt
from keras.layers import Input, Dense, Conv2D, MaxPool2D,
GlobalAveragePooling2D
from keras.models import Sequential, Model
from sklearn.metrics import accuracy_score, classification_report,
confusion_matrix
import tensorflow as tf
from google.colab import drive
drive.mount('/content/drive')
import tensorflow as tf
import os
import random
from tensorflow.keras.preprocessing import image_dataset_from_directory
from sklearn.utils import class_weight
import numpy as np
import shutil
from google.colab import drive
drive.mount('/content/drive', force_remount=True)
augmented_dir = '/content/drive/MyDrive/augmented_data'
base_split_dir = '/content/drive/MyDrive/augmented_split'
# Folders
train_dir = os.path.join(base_split_dir, 'train')
valid_dir = os.path.join(base_split_dir, 'valid')
test_dir  = os.path.join(base_split_dir, 'test')
# Create directories if not exist
for folder in [train_dir, valid_dir, test_dir]:
    os.makedirs(os.path.join(folder, 'anemic_augmented'), exist_ok=True)
    os.makedirs(os.path.join(folder, 'non_anemic_augmented'),
exist_ok=True)


def split_files(src_dir, train_dir, valid_dir, test_dir, train_ratio=0.7,
valid_ratio=0.2):
    files = [f for f in os.listdir(src_dir) if
os.path.isfile(os.path.join(src_dir, f))]
    random.shuffle(files)
    n = len(files)
    n_train = int(n * train_ratio)
    n_valid = int(n * valid_ratio)

    for i, file in enumerate(files):
        src_file = os.path.join(src_dir, file)
        if i < n_train:
            dst = os.path.join(train_dir, file)
        elif i < n_train + n_valid:
            dst = os.path.join(valid_dir, file)
        else:
            dst = os.path.join(test_dir, file)
        shutil.copy(src_file, dst)

  # Split classes
split_files(os.path.join(augmented_dir, 'anemic_augmented'),
            os.path.join(train_dir, 'anemic_augmented'),
            os.path.join(valid_dir, 'anemic_augmented'),
```

```
            os.path.join(test_dir,  'anemic_augmented'))

split_files(os.path.join(augmented_dir, 'non_anemic_augmented'),
            os.path.join(train_dir, 'non_anemic_augmented'),
            os.path.join(valid_dir, 'non_anemic_augmented'),
            os.path.join(test_dir,  'non_anemic_augmented'))

print("Data splitting completed!")

IMAGE_SIZE = 224
BATCH_SIZE = 16

train_dataset = image_dataset_from_directory(
    train_dir,
    image_size=(IMAGE_SIZE, IMAGE_SIZE),
    batch_size=BATCH_SIZE,
    label_mode='int',
    shuffle=True
)

valid_dataset = image_dataset_from_directory(
    valid_dir,
    image_size=(IMAGE_SIZE, IMAGE_SIZE),
    batch_size=BATCH_SIZE,
    label_mode='int',
    shuffle=False
)
test_dataset = image_dataset_from_directory(
    test_dir,
    image_size=(IMAGE_SIZE, IMAGE_SIZE),
    batch_size=BATCH_SIZE,
    label_mode='int',
    shuffle=False
)
import tensorflow as tf

import tensorflow as tf

# Custom function for 90Â°/270Â° rotations and vertical flips
def custom_augmentation(image):
    # Randomly apply 90Â° or 270Â° rotation
    k = tf.random.uniform(shape=[], minval=0, maxval=2, dtype=tf.int32)
    if k == 0:
        image = tf.image.rot90(image, k=1)  # 90Â°
    else:
        image = tf.image.rot90(image, k=3)  # 270Â°

    # Randomly flip vertically
    image = tf.image.random_flip_up_down(image)

    return image

# Combine with standard Keras augmentations
data_augmentation = tf.keras.Sequential([
    tf.keras.layers.Lambda(custom_augmentation),  # Apply custom
90Â°/270Â° rotations & vertical flip
    tf.keras.layers.RandomFlip("horizontal"),     # Random horizontal
flip
```

```
    tf.keras.layers.RandomRotation(0.04),         # Random small rotation
Â±4%
    tf.keras.layers.RandomZoom(height_factor=(-0.05, 0.05),
width_factor=(-0.05, 0.05)),
    tf.keras.layers.RandomTranslation(height_factor=0.05,
width_factor=0.05),
])

def augment(images, labels):
    return data_augmentation(images, training=True), labels

train_dataset = train_dataset.map(augment,
num_parallel_calls=tf.data.AUTOTUNE);
train_dataset = train_dataset.prefetch(tf.data.AUTOTUNE)
valid_dataset = valid_dataset.prefetch(tf.data.AUTOTUNE)
test_dataset = test_dataset.prefetch(tf.data.AUTOTUNE)
# Extract labels from train_dataset
train_labels = np.concatenate([y.numpy() for x, y in train_dataset],
axis=0)
class_weights = class_weight.compute_class_weight(
    class_weight='balanced',
    classes=np.unique(train_labels),
    y=train_labels
)
class_weights_dict = {i: w for i, w in enumerate(class_weights)}
print("Class weights:", class_weights_dict)

#CNN Model
from tensorflow.keras import layers
from tensorflow.keras.layers import Input, Conv2D, MaxPool2D,
GlobalAveragePooling2D
from tensorflow.keras.layers import Dense, BatchNormalization, Dropout
from tensorflow.keras.models import Model
from keras.models import Sequential, Model
from keras.layers import Input, Dense, Conv2D, MaxPool2D,
GlobalAveragePooling2D
inputs = Input(shape=(IMAGE_SIZE, IMAGE_SIZE, 3))


inputs = Input(shape=(224, 224, 3))

x = Conv2D(32, (3,3), padding="same", activation="relu")(inputs)
x = BatchNormalization()(x)
x = MaxPool2D(2,2)(x)

x = Conv2D(64, (3,3), padding="same", activation="relu")(x)
x = BatchNormalization()(x)
x = MaxPool2D(2,2)(x)

x = Conv2D(128, (3,3), padding="same", activation="relu")(x)
x = BatchNormalization()(x)
x = MaxPool2D(2,2)(x)

x = Conv2D(256, (3,3), padding="same", activation="relu")(x)
x = BatchNormalization()(x)

x = GlobalAveragePooling2D()(x)
x = Dense(128, activation="relu")(x)
```

```
x = Dropout(0.4)(x)

outputs = Dense(1, activation="sigmoid")(x)

model = Model(inputs, outputs)
model.summary()

model.compile(
    loss=tf.keras.losses.BinaryCrossentropy() ,
    optimizer=tf.keras.optimizers.Adam(),
    metrics=['accuracy']
)
history = model.fit(train_dataset, validation_data=valid_dataset,
epochs=100, class_weight=class_weights_dict)
model.save('model_anemia.h5')
"""VISULIZE MODEL """
plt.plot(np.arange(100) , model_information['loss'] , label="loss")
plt.plot(np.arange(100) , model_information['val_loss'] ,
label="val_loss")
plt.title('MODEL LOSS AND VALIDATION LOSS')
plt.savefig('MOBILE_pretrained_loss.png')
plt.legend()
plt.show

"""VISULIZE MODEL """
plt.plot(np.arange(100) , model_information['accuracy'] ,
label="accuracy")
plt.plot(np.arange(100) , model_information['val_accuracy'] ,
label="val_accuracy")
plt.title('MODEL accuracy AND val_accuracy LOSS')
plt.savefig('MOBILE_pretrained_val_accuracy.png')
plt.legend()
plt.show()

import os

# Correcting the path to the test data directory
test_data_path = test_dir # Using the previously defined test_dir
test_images = []

for label_name in os.listdir(test_data_path):
    class_dir = os.path.join(test_data_path, label_name)
    if os.path.isdir(class_dir):
        for img in os.listdir(class_dir):
            img_path = os.path.join(class_dir, img)
            test_images.append([img_path, label_name])

print("Number of test images:", len(test_images))
print(test_images[:5])

import random
random.shuffle(test_images)

test = test_images[0]
img = test[0]
label = test[1]

class_names = train_generator.class_indices
```

```
classes = list(class_names.keys())
classes

model.evaluate(test_dataset)


#TEST VISULIZATION
from tensorflow.keras.preprocessing import image
def preprocess_image(img_path):
    img = image.load_img(img_path, target_size=(224, 224))  # Assuming
your model expects 64x64 images
    img = image.img_to_array(img)
    img = img /255.0
    img = np.expand_dims(img, axis=0)
    return img

# Function to predict class and visualize the image
def predict_and_visualize(image_paths):
    plt.figure(figsize=(10, 10))
    for i, img_path in enumerate(image_paths):
        img = preprocess_image(img_path[0])
        prediction = model.predict(img , verbose=0)

        predicted_class = classes[int(prediction.round()[0][0])]

        # Load and display the image
        plt.subplot(4, 5, i+1)
        plt.imshow(image.load_img(img_path[0]))
        plt.title(f' real: {img_path[1]} \n Predicted: {predicted_class}'
, fontsize=10 )
        plt.axis('off')
    plt.savefig('result.png')
    plt.show()

predict_and_visualize(test_images[:14])

@Author : Fatemeh_Kn
```
